# Hidden Parameters Impacting Resurgence of SARS-CoV-2 Pandemic^*^

**DOI:** 10.1101/2021.01.15.20248217

**Authors:** Yi Zhang, Sanjiv Kapoor

## Abstract

The spread of the COVID-19 virus has had an enormous impact on the world’s health and socioeconomic system. While lockdowns, which severely limit the movement of the population, have been implemented in March 2020 and again recently, the psychological and economic cost are severe. Removal of these restrictions occurred with varying degrees of success.

To study the resurgence of the virus in some communities we consider an epidemiological model, SIR-SD-L, that incorporates introduction of new population due to the removal of lockdown and identify parameters that impact the spread of the virus. This compartmental model of the epidemic incorporates a social distance metric based on progression of the infections; it models the dynamic propensity of infection spread based on the current infections relative to the susceptible population. The model is validated using data on growth of infections, hospitalizations and death, considering 24 counties in multiple US states and a categorization of the lockdown removal policies after the first lockdown. Model parameters, which include a compartment for the isolated population, are used to determine the rate at which the susceptible population increases to fit the rate of infections. Along with social distancing mandates, we identify active infections and the susceptible population as important factors in the resurgence of infections. We measure the efficacy of the lockdown removal policy via a ratio, PIR, which evaluates to less than 1 for counties where social distancing measures were mandated and which delayed complete re-opening of closed spaces like bars and restaurants. For other counties this ratio is greater than 1.

We also studied infection growth in the 24 US counties with respect to a release policy derived from CDC guidelines and compared against strategies that delay the removal of lockdown.

Our results illustrate that guidelines for deciding when lockdown rules are to be relaxed should consider the current state of the infectious population and the remaining susceptible population, hidden parameters that are deducible from models such as SIR-SD-L, and not limit policy considerations to the rate of new infections alone. This is especially true for counties where the growth rate of the virus is initially slow and misleading. Emphasis on social distancing is critical.

## 1 Introduction

The current health crisis induced by the SARS-CoV-2 virus has resulted in lockdown procedures in countries around the world and is also marked by considerable debate over the use of social distancing, including the use of masks. While guidelines have been issued, adherence has been patchy.

In this context, timing the removal of lockdown and social distancing guidelines play an important role in controlling the spread and impact of the COVID-19 pandemic (*1*). Since the incubation period of the disease results in delayed impact on the progression of the disease, long term accurate modeling of the spread of the virus is critical. While it is anticipated that lockdown removal will result in resurgence of the virus, the impact of the policies are not clear on the resurgence. In some countries that adopted stringent controls when the lockdown was removed, e.g. NewZealand and Thailand (*2*), there was considerable success in controlling the virus. In other countries there was considerably less success (*3*).

An analysis of the spread of infection after the lockdown ends is critical so as to determine policies that could be used to prevent capacity issues in the healthcare system (*4*). The resurgence in some states of US in the months following the removal of the first lockdown has been especially concerning. We construct a model to account for this resurgence utilizing a release mechanism incorporated into a compartmental model and identify key factors that influence the resurgence of the virus. We consider 24 counties in US and illustrate the importance of determining the number of active infections as well as the susceptible population left when planning for removal of lockdown. Government mandated policies also played a substantial role.

The analysis is based on epidemic spread models, adapted from the SIR model (*5, 6*) (originally from Kermack and McKendrick (*7–9*)). A key differentiation in this work is the use of dynamic social distancing factor in the model, termed the SIR-SD model. Prior evidence that the reduction in social contacts has consequent impact on the reproduction number *R*_0_ has been investigated in (*10*). The methodology requires the use of matrices that model the contact between population groups and estimates of the matrix may be difficult to obtain. The contact patterns assumed are, however, not dynamic. Dynamic changes in the transmission factor of the infection as a function of government social distancing measures, has also been modeled in (*11*). Additionally, prior research on the impact of social distancing has been considered by Kissler et al. (*12*) which considers the impact of intermittent social distancing on critical care capacity. Recent modeling efforts have focused on specific covariates like mobility (*13*) and modeling and analysis of various other aspects of the COVID-19 pandemic is widespread (*14–22*).

In this research, we consider an alternate viewpoint where, in this model, we use a social distancing factor to modify the virus transmission rate that reflects the behavior of the population as the number of infection cases rises and reporting of the infections, as well as government mandates, becomes more prevalent. The transmission of the virus incorporates a factor that is inversely dependent on the number of cumulative infections and that incorporates a time-dependent susceptible population size.

We focus our attention on select counties in US that showed large number of infections. Our analysis of the time period between March and September, 2020, illustrates that removal of lockdown leads to increased rates of infection, especially in counties that had fewer mandates on social distancing measures and allowed closed spaces, including bars and restaurants, to reopen. The analysis utilizes a ratio, *PIR*, which measures the peak daily new infections after removal of lockdown relative to the peak daily new infections during the lockdown period. This ratio is related to the ratio of active infections at reopen relative to the peak active infections during lockdown. Additionally the ratio, *PIR*, increases with the percentage of susceptible population left. Statistics of these ratios are not accurately known but these hidden parameters can be estimated from the model we propose. In our analysis, the ratio *PIR* was less than one for counties with strong social distancing mandates and was much greater than one for other counties.

The model we develop is also used to compare the criteria for reopening. We compare the cumulative new cases that resulted from the actual reopening time against a two-phase downward trajectory policy that emerges from CDC guidelines (*23*) as well as against a policy that further delayed reopening by 1 month (30 days). The two-phased trajectory only accounts for the current rate of infection and does not account for factors that have shown to be important in our analysis, the susceptible population left and the current count of the infected population. The results illustrate the benefits of delayed reopening in terms of case counts, as this reduces the active infections in the community.

In summary, we advocate the use of epidemiology models that can accurately estimate the number of active infections and the susceptible population to guide the re-open policy. The rate at which population is released from lockdown also impacts the resurgence and its impact should be computed from the model. Furthermore, there is a stark contrast between counties that used social distancing measures like strict usage of masks and restricted use of crowded spaces and those that did not have restrictions.

Publicly available data from multiple repositories (*24–26*) was used.

## 2 Viral Resurgence

Guidelines and timing of easing of lockdown conditions are very critical, requiring an analysis of the pandemic growth rate, especially when lockdown has been applied early and the impact of the virus is less evident. Policy based on limited local spread of the virus can be misleading. To identify the impact of the number of active infections, the number of susceptible people left and the population size on the resurgence of the virus, the analysis presented below, utilizes a model that incorporates infections, hospitalizations as well as the population under lockdown.

### 2.1 SIR-SD-L Model

Our model incorporates population compartments that include the susceptible population *S*, the possible hosts to the virus, the currently infected population *I*, the set of population that are hospitalized *I*_*H*_ and those that are tested and non-hospitalized set *I*_*N*_. *R* is the recovered population and *D* the dead. The impact of the lockdown is via the population that is in the set marked *H. H* is the population that went into effective lockdown with the the assumption that no interaction of this population exists with the infected population. On removal of lockdown, a fraction of the population transitions from *H* to the set *S*. As compared to the standard SEIR model we eliminate the transition to the exposed set as data for this transition has to be inferred or adopted from literature and is not useful in our model. We also do not have an explicit asymptomatic population; we assume that the impact of this is implicit in the infection rate. Instead we allow the population to transit directly from *S* to *I* with the assumption that the susceptible fraction is *ηN*, to account for the fraction that may form a host to the virus at a point of time. Furthermore the testing and reporting of infections include some of the asymptomatic cases and all of this population is in the *I* with transitions leading to hospitalizations or otherwise. In addition to the compartments in Figure 1, we also keep track of the cumulative cases *G*.

**Figure 1:**
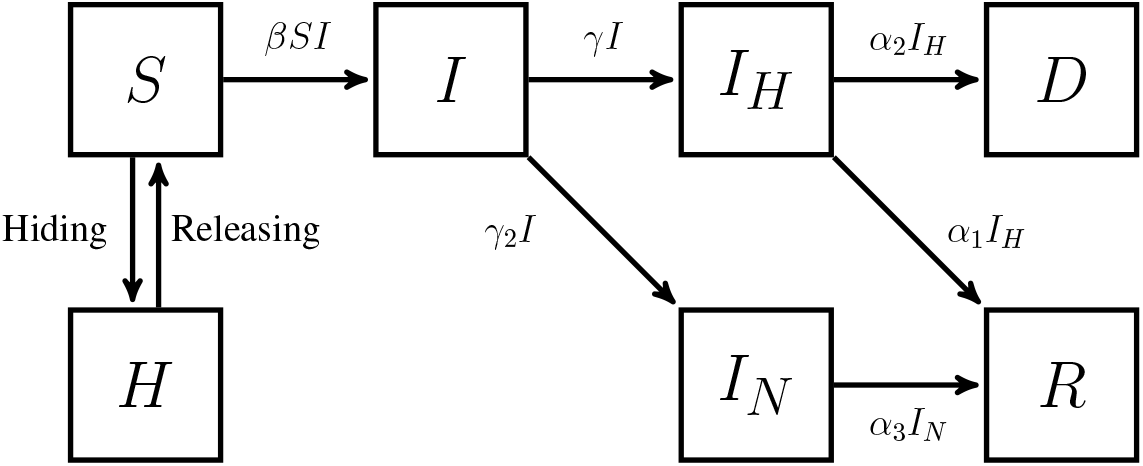
The SIR-SD-L Compartment Mode

Changes over time in the population categories are modeled by the differential equations:

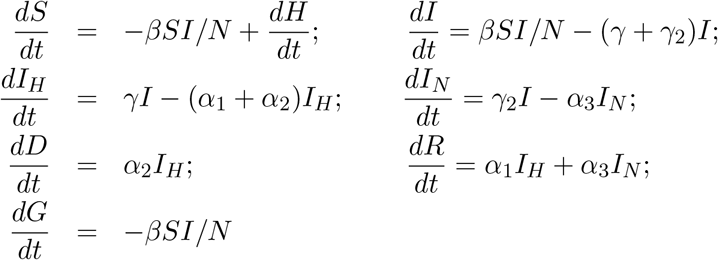

where *β* is the infection rate and *γ* is the rate of transition from infected set *I* to *I*_*H*_. *γ*_2_ is the transition rate from *I* to *I*_*N*_. The rates *α*_1_, *α*_2_ and *α*_3_ are transitions between compartments as illustrated in Figure 1. *H* is a time-varying function that represents the population that is under “lockdown”.

For the purpose of our analysis we consider that *dH/dt* is a constant, corresponding to a linear release function; hence the term L in the model name SIR-SD-L.

#### Social Distancing impact

To model the impact of social distancing, we consider a modification of the SIR model with social distancing where *β*, the infection rate, is dynamic. In this model, *β* is dependent on the infection spread as measured by the number of cumulative infections. While retaining the other properties of the SIR model, we let the function *β* be defined as:

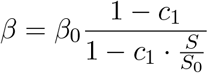

where *S*_0_ = *ηN* is the initial susceptible population and *β*_0_ a constant. When additional population is released from *H*, the parameter *β* becomes:

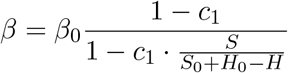

where *H*_0_ is the initial population under lockdown and *H* the current population under lockdown. This model measures the psychological “fear” impact of of the spread of the virus. Other models have studied impact of covariates that include mobility, interventions, density etc. on the variable *β* via neural networks (*13*). Incorporating the social distancing model provides for a better fit to the infection dynamics. We illustrate this on the standard SIR model during the period of lockdown corresponding to each county. The actual dates differed across counties. We choose 24 counties in the US to fit the model, with the model parameters being estimated by fitting to available data on cumulative infections, hospitalizations and deaths. The loss function is discussed in the appendix and here we compare the two models using weighted Root Mean Squared Error (RMSE) to quantify the error in estimating the cumulative infection data where the weighting geometrically decreases, starting at the end of the time series. The fitting is illustrated for a few counties in Figure 2.

**Figure 2:**
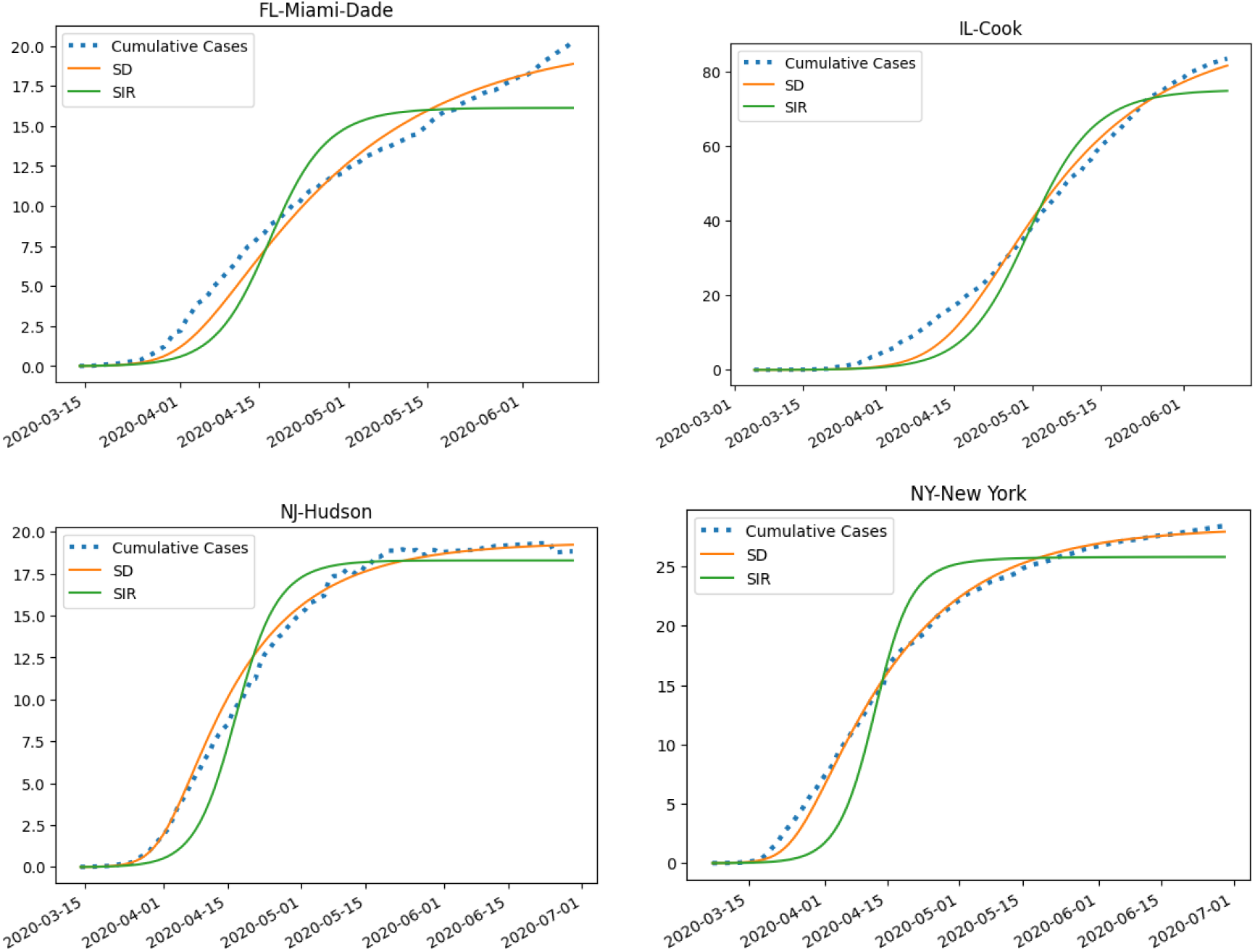
Graphs illustrating the fitting comparison between SIR-SD model with social distancing (SD) and the standard SIR model for cumulative infection cases Miami-Dade, Cook, Hudson and New York counties in US. See additional counties in the appendix.

The weighted Root Mean Squared Error of the cumulative cases is provided in Table 1 to show the comparison between the two models. For some of counties in New Jersey and New York we see substantial improvement of RMSE values. On average the SIR-SD model reduces the RMSE by 28% as compared to the standard model SIR model.

**Table 1:** Table of weighted RMSE of cumulative cases for counties around USA illustrating fit of SIR-SD model in comparison to the standard SIR model. Refer to Appendix for graphs.

| State - County | SIR-SD | SIR | State - County | SIR-SD | SIR |
| --- | --- | --- | --- | --- | --- |
| AZ-Maricopa | 601.6481 | 827.7992 | NJ-Bergen | 282.0097 | 1261.5664 |
| CA-Los Angeles | 4095.9119 | 6084.3161 | NJ-Hudson | 567.5651 | 1341.2966 |
| CA-Riverside | 901.6782 | 1054.0520 | NV-Clark | 377.1504 | 652.4627 |
| FL-Broward | 779.9327 | 977.0944 | NY-New York | 507.3686 | 2647.4040 |
| FL-Miami-Dade | 857.6028 | 1933.3845 | OH-Franklin | 255.3531 | 294.9549 |
| GA-Fulton | 310.8251 | 425.9091 | PA-Philadelphia | 1058.7648 | 1952.0696 |
| IL-Cook | 3057.1101 | 5761.1191 | TN-Shelby | 646.7721 | 701.5321 |
| LA-Jefferson | 453.9668 | 540.9758 | TX-Dallas | 495.7981 | 550.3643 |
| MA-Middlesex | 370.0816 | 1236.3973 | TX-Harris | 1019.9023 | 1350.4305 |
| MD-Prince George's | 1357.9088 | 1063.8959 | UT-Salt Lake | 157.4909 | 198.7106 |
| MN-Hennepin | 189.2487 | 120.5044 | VA-Fairfax | 559.2513 | 538.3087 |
| NC-Mecklenburg | 333.5950 | 355.7503 | WI-Milwaukee | 709.9368 | 882.3074 |

### 2.2 Impact of Removal of lockdown population

After the relative success of the initial lockdown a phased approach was specified which would have moved states to various stages of normalcy. Unfortunately this resulted in resurgence of the virus in multiple counties and states. Each of the individual states had defined their own policies based on CDC guidelines and that resulted in removal of lockdown with somewhat different policies and mandates. While many states did not mandate masks and additionally had few restrictions mandated for places where large gatherings might take place, other states had restrictions which were eased very cautiously.

Data was obtained about the announced reopen dates of each county from news and government website (*24, 27*). Also recorded were the policies on face covering which were categorised into 3 color-coded categories. Counties which explicitly emphasized the mandated restrictions including requiring face covering all the time in public places and had strict guidelines on bars and restaurants were colored red. Counties with milder emphasis and recommendations on face coverings, either by the state or by counties themselves were allocated yellow. A green color meant that there was no explicit mandate announced separate from CDC guidelines. The details can be found in Table 2. State government guidelines were used when no specific county guideline was available.

**Table 2:** Dates and Policy guidelines for removal of lockdown. Green color indicates no specific mandate for masks and restrictions during reopening while red color indicates face mandates and restrictive openings. Yellow indicates mild mandates on masks and reopening.

| County | Color | Date | County | Color | Date |
| --- | --- | --- | --- | --- | --- |
| AZ-Maricopa | green | 2020-05-28 | NJ-Bergen | red | 2020-06-22 |
| CA-Los Angeles | yellow | 2020-06-12 | NJ-Hudson | red | 2020-06-22 |
| CA-Riverside | yellow | 2020-06-12 | NV-Clark | green | 2020-05-29 |
| FL-Broward | green | 2020-06-12 | NY-New York | red | 2020-06-22 |
| FL-Miami-Dade | green | 2020-06-03 | OH-Franklin | green | 2020-05-21 |
| GA-Fulton | green | 2020-06-12 | PA-Philadelphia | yellow | 2020-06-05 |
| IL-Cook | red | 2020-06-03 | TN-Shelby | green | 2020-06-15 |
| LA-Jefferson | green | 2020-06-05 | TX-Dallas | green | 2020-05-22 |
| MA-Middlesex | red | 2020-06-22 | TX-Harris-Houston | green | 2020-06-03 |
| MD-Prince Georges | red | 2020-06-29 | UT-Salt Lake | yellow | 2020-05-15 |
| MN-Hennepin | green | 2020-06-04 | VA-Fairfax | red | 2020-06-12 |
| NC-Mecklenburg | green | 2020-05-22 | WI-Milwaukee | yellow | 2020-07-01 |

To model the release from lockdown via the proposed linear release function, we added population back from the population set *H* to the susceptible on a daily basis, until an estimated number of people, *H*_0_ have emerged from lockdown. The release started 7 days after the date a county enters phase 2 of reopening; this models an initial phase of cautious reopening. *H*_0_ is obtained by the algorithm that fits the model. Due to change in the demography of the population newly added to the susceptible pool, the reopen phase introduces 2 new parameters, *k, k*_2_. The rate of release is represented by the parameter *h*, the fraction of total population under lockdown that is released everyday into the susceptible set, *i*.*e. h* = 0.05 implies that the estimated population *H*_0_ that emerges from lockdown is released linearly in 20 days. *k* and *k*_2_ are multipliers for *γ* and *α*_2_, respectively, in the reopen phase. We observed a decrease in hospitalization and mortality rates as different counties entered the reopen phase and use these 2 new parameter to account for that change. The parameter estimation outcome for 24 different counties is in Figure 3, which illustrates the graph of the cumulative infections obtained from fitting our model, as well as the predicted count of deaths. The shaded region in each figure illustrates the range of predictions with 95% confidence interval (details of the computation are in the appendix). The actual data is mostly contained within the confidence interval bands provided by the model predictions.

**Figure 3:**
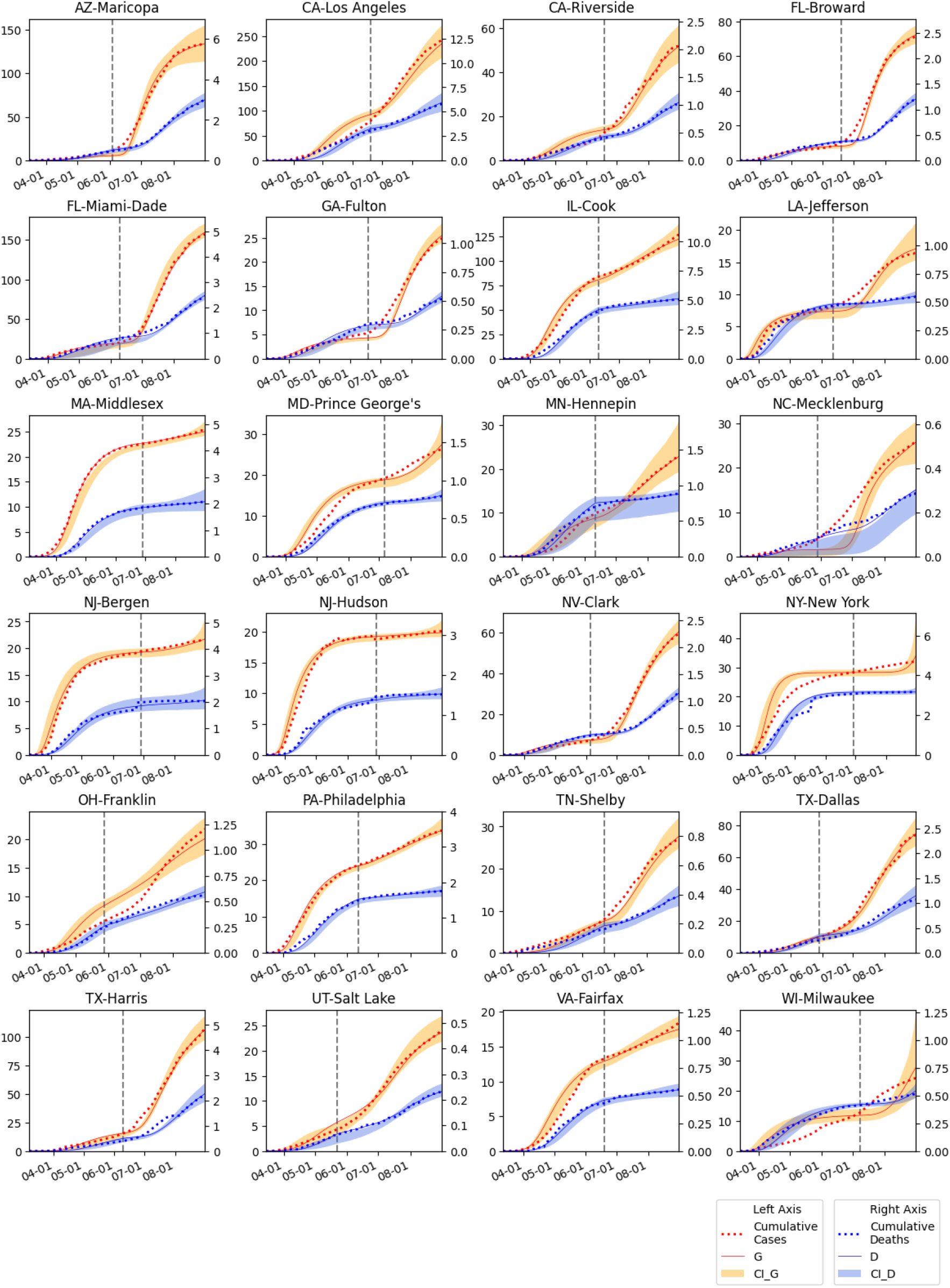
The SIR-SD-L model fitted for 24 US counties. Cumulative infection numbers (G) and deaths (D) are modeled for each of the counties. Range of intervals (95% CI) are illustrated for both G and D. The vertical dotted line represents the reopen date for each county.

To measure the parameters that impact the reopening or removal of lockdown, we investigated the peak infection ratio, *PIR*, for each county during the time frame mid-March to end of August. *PIR* is the ratio of the peak daily new cases after reopen relative to the peak daily new case before reopen during this time frame. We correlate this with *PAR*, the peak active ratio as determined by *I*_*reopen*_*/I*_*peak*_ which is the ratio of the active cases when a county enters reopen phase relative to the peak active cases before reopen. We then correlate *PIR* with *SLR* = *S*_*reopen*_*/S*_0_, the ratio of susceptibles at reopen with respect to the initial susceptible population. We also correlate *PIR* with *HSR* = *H*_0_*/S*_0_ the ratio of the population that emerges from lockdown over the initial exposed susceptible population. An increased value of *H*_0_ could represent the set of people that are taking additional risks after the removal of lockdown. An interesting relationship emerges between log(*PIR*) and *SLR* and *HSR* (Figure 4), As the figures suggest, slower release of the population guided by mandates and emphasis on social distancing policies that include masks as well as limitations on public gatherings, influence the ratio, *PIR*, and help limit the next peak. This is shown by the increasing relation between the log of the peak ratio, log(*PIR*) and the other ratios, *HSR, SLR* and *PAR*. Interestingly for counties that are colored red, log(*PIR*) is negative while log(*PIR*) is positive for counties colored green. This illustrates the impact of state and county policies on the resurgence of the virus. Counties that had lower *PIR* had either less active cases or less susceptible population at the time of reopening, further indicating the importance of timing the reopening.

**Figure 4:**
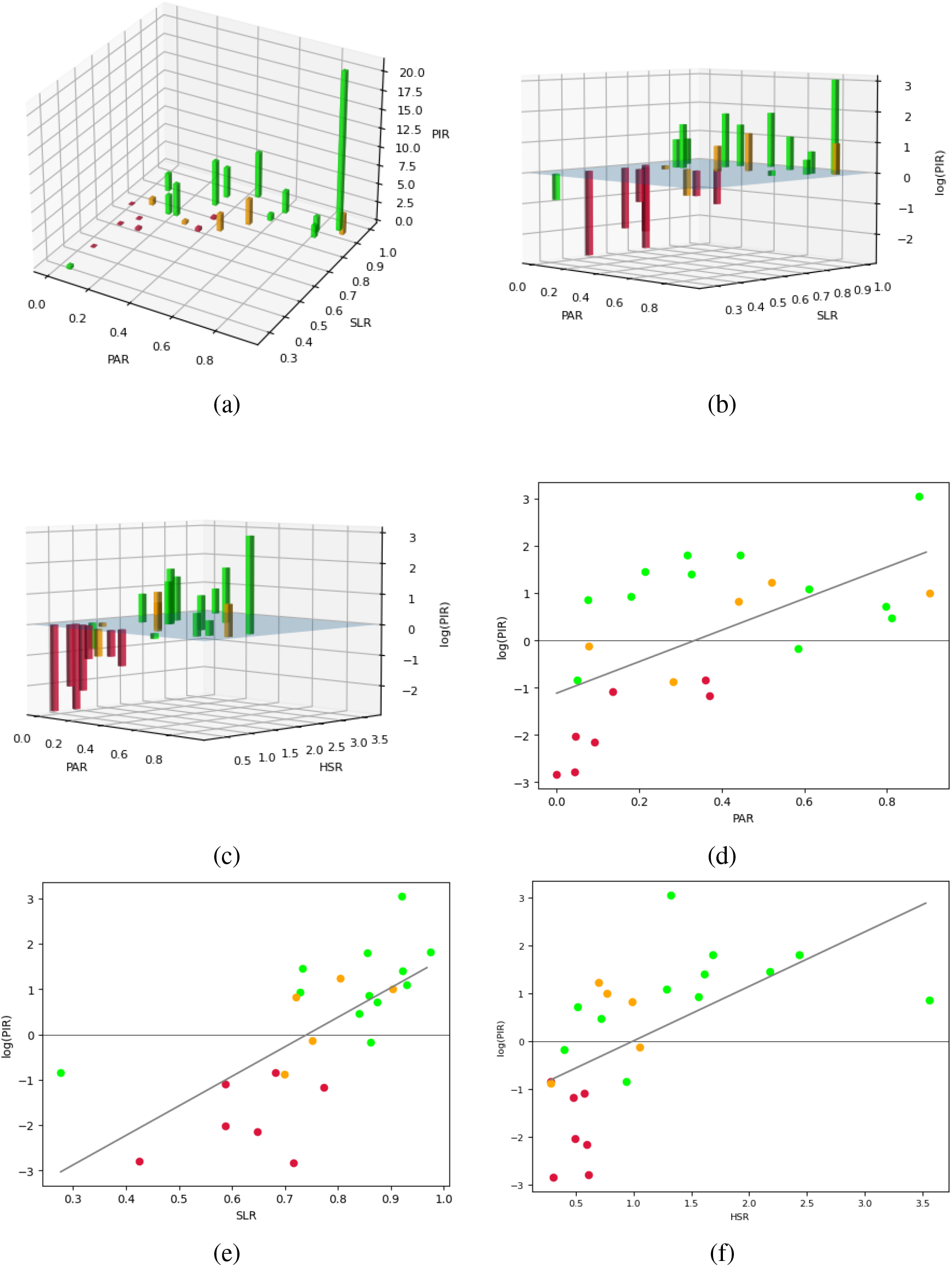
(a) Daily peak infections ratio *PIR* vs *PAR*(*I*_*reopen*_*/I*_*peak*_), and *SLR*(*S*_*reopen*_*/S*_0_). (b) log(*PIR*) vs *PAR*(*I*_*reopen*_*/I*_*peak*_) and *SLR*(*S*_*left*_*/S*_0_). (c) log(*PIR*) vs *PAR* and *HSR*(*H*_0_*/S*_0_). (d),(e), (f) plots of log(*PIR*) vs each of *PAR, SLR* and *HSR*.

## 3 The impact of delayed release

Timing the reopening, an important aspect of the resurgence, was primarily based on government guidelines. The CDC (*23*) recommendations led to a federal plan for re-opening after lockdown. The plan was published in a document outlining conditions for phased re-opening in mid April (*28*), which specifies the threshold for entering successive reopen phases. From these guidelines we extracted the primary rule where entry into each of two phases, phase 2 and phase 3 required a downward trajectory of new cases for 14 days, with some allowances. We implemented these rules to establish a time for reopening into phase 3 for each of the counties. We term this the Two-phase Rule and compare this rule against the schedule for actual removal of lockdown that occurred, as well as against rules that delayed the release of lockdown further by a week to a month.

To illustrate the impact of the opening strategies we consider 6 counties. where the two phases were almost consecutive in that there was a consistent decrease of new active cases (Figure 5). For multiple other counties, the appendix illustrates the time of opening into phase 3 based on the Two-Phase rule. For each of the counties, the blue curve, in the left part of the corresponding figure, is a cubic spline smoothing (with coefficient 0.1) on the 14-day moving average of new cases. The 2 green time windows are the detected 14-day downward trajectories, in which we allow 3 days of increase as a grace period, following the CDC guidelines. After the detection of the 2 downward trajectories (decreasing new case counts) a county was progressed into phase 3 reopening in accordance with the Two-phase rule. The vertical dotted line is the actual date the county entered phase 3 as indicated from government sources. In the figures to the right we illustrate simulated cumulative cases when lockdown was released according to multiple policies. The shaded area is the cumulative infection following the actual removal of lockdown while *G*_*CDC*_ illustrates the growth when a county starts phase 3 following the Two-phase guideline. We also simulate the scenarios when the actual reopening is delayed by 1 week and 1 month (30 days). All the simulations utilized the model developed earlier in the paper.

**Figure 5:**
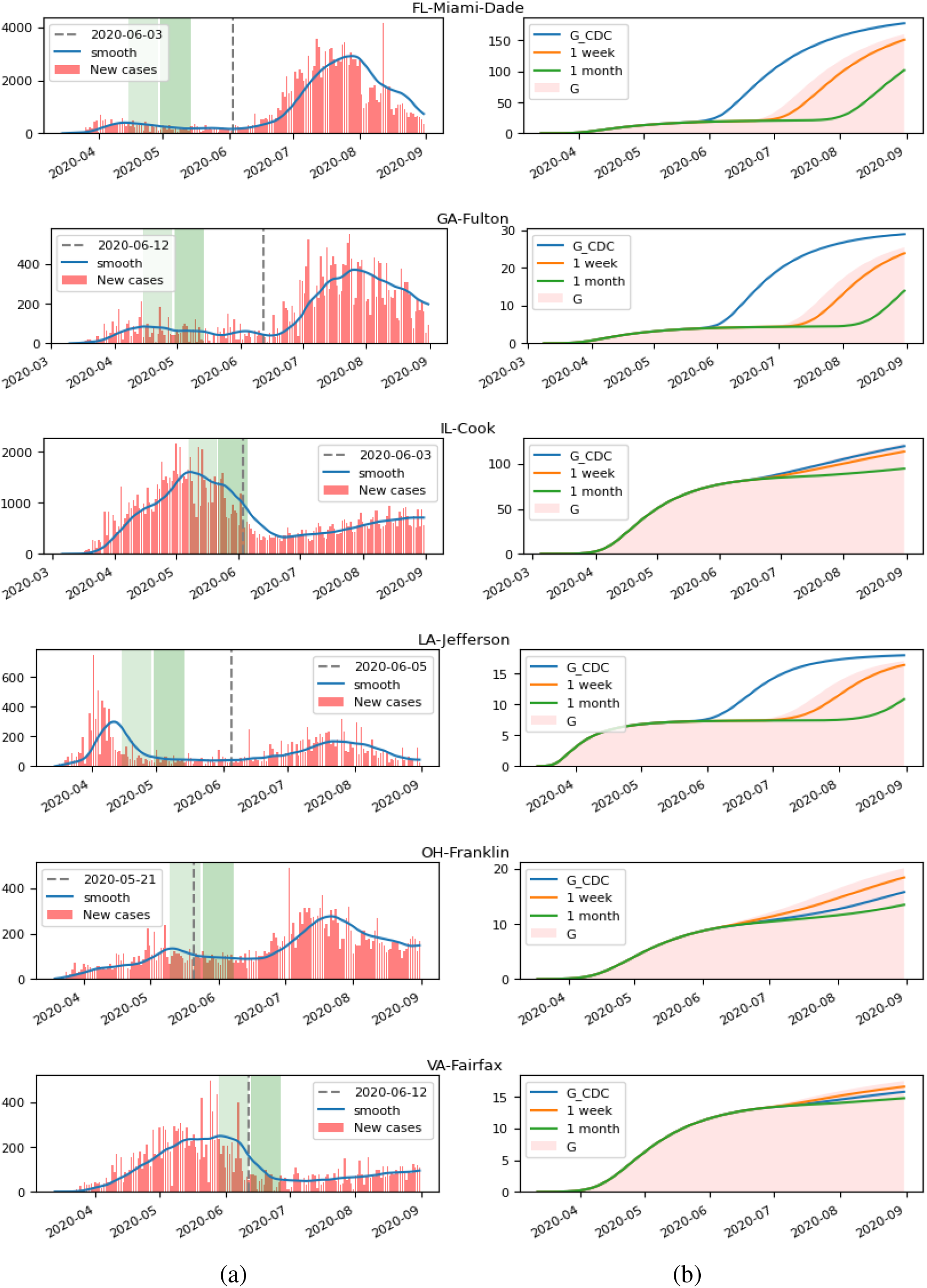
(a) *G*_*CDC*_: Re-opening via the Two-phase rule (each phase of 14 days represented by the green bands) as compared to actual re-open date (dashed vertical line). The blue curve represents 14-day moving average of the actual daily cases. (b) Simulated comparison in cumulative infection count for actual reopening as compared to reopening following *G*_*CDC*_ guidelines and to delayed opening after 1 week and 1 month delay.

As illustrated in Figure 6, delaying the opening would have helped in reducing the total number of the cases. While delaying by 1 week reduced the number of infections, Table 3 illustrates that delaying by 1 month could have resulted in substantial less infections in some counties. The delay ratio is the ratio of the new infections after reopen when the reopen is delayed by 1 month, versus the actual new infection cases after the actual reopening date. An average reduction of new cases by 42% is illustrated within the time frame we considered. Over an extended period of time the improvement would likely be less.

**Table 3:** Table of delay ratios of new infection counts if reopening is delayed for 1 month.

| State - County | Delay Ratio | State - County | Delay Ratio |
| --- | --- | --- | --- |
| AZ-Maricopa | 0.955836 | NJ-Bergen | 0.151275 |
| CA-Los Angeles | 0.550668 | NJ-Hudson | 0.307536 |
| CA-Riverside | 0.717784 | NV-Clark | 0.793129 |
| FL-Broward | 0.908649 | NY-New York | 0.00462 |
| FL-Miami-Dade | 0.839463 | OH-Franklin | 0.558942 |
| GA-Fulton | 0.826248 | PA-Philadelphia | 0.373862 |
| IL-Cook | 0.416732 | TN-Shelby | 0.636687 |
| LA-Jefferson | 0.834068 | TX-Dallas | 0.790467 |
| MA-Middlesex | 0.26935 | TX-Harris | 0.770216 |
| MD-Prince George's | 0.218844 | UT-Salt Lake | 0.762163 |
| MN-Hennepin | 0.521526 | VA-Fairfax | 0.457507 |
| NC-Mecklenburg | 0.935439 | WI-Milwaukee | 0.305277 |

**Figure 6:**
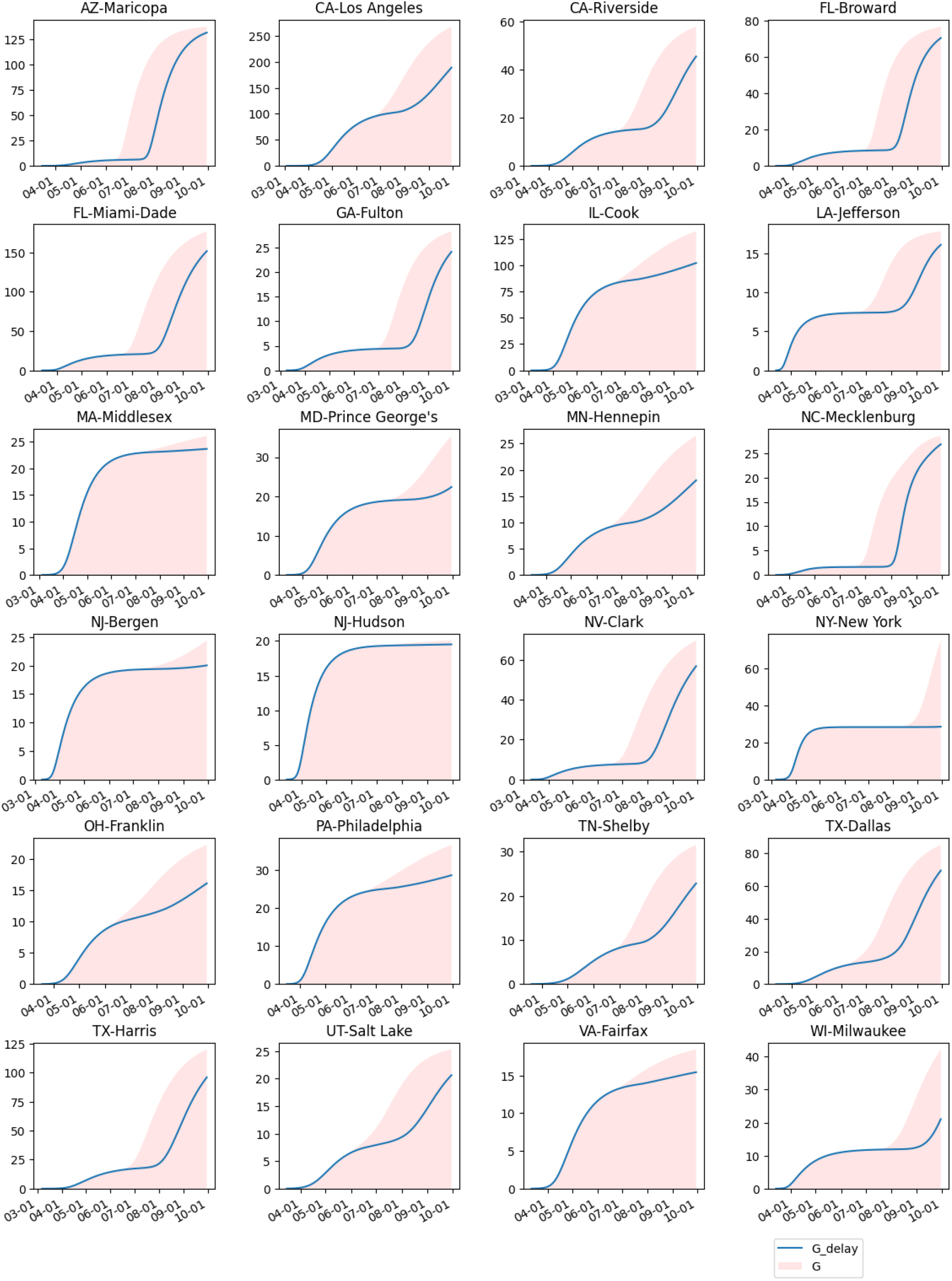
Comparison of cumulative cases with delayed reopening (delayed by 1 month) simulation represented by blue curve

## Data Availability

Data from public sources was used and referenced.

https://github.com/CSSEGISandData/COVID-19.

## 4 Acknowledgements

We acknowledge the use of cloud computing resources provided by the Chameleon project at Chameleon-cloud.org.

### 5 Appendix: Supplementary Materials

#### 5.1 Growth rate of Infection

We model the growth rate of the infection compartments by the system of differential equation below:

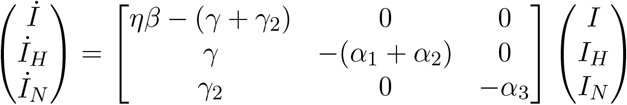

The eigenvalues of the matrix characterizing the linear system are

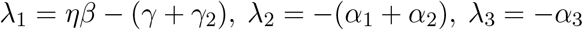

and the corresponding eigenvectors are

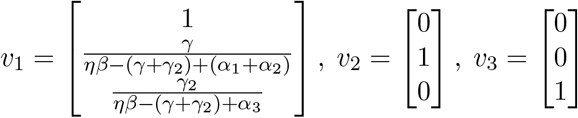

Thus,

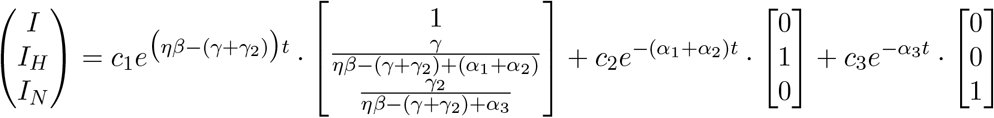

The value of *R*_0_ is obtained from transition analysis as:

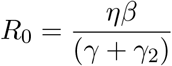

#### 5.2 Methods

We use a discrete-step version to compute the model parameters. In all the models we start the fitting on the day when there are more than 5 reported cases and stop at the end of August to reduce errors due to long term impact of reopening. We tried two standard constrained optimization methods “L-BFGS-B” and “TNC” but did not observe much difference between the two.

Model parameters were obtained using a program to fit the differential equations. The variables used in the model are *η, β, γ, γ*_2_, *α*_1_, *α*_2_, *α*_3_, *c*_1_, where *η* specifies the susceptible set, i.e. *S*_0_ = *ηN*. An optimizer was used to fit determine the parameters using a loss function

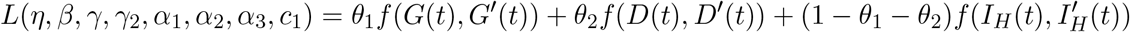

where *G* (*t*) is the computed cumulative infections, *G*(*t*) is the actual data time series of cumulative infected cases; similarly *D*(*t*) (*I*_*H*_(*t*)) and 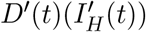 represent the cumulative deaths (active hospitalization) in the model and from the actual data, respectively. The function *f* was chosen to minimize the RMSE, alternatively maximize the R^2^ value (coefficient of determination) of the data with a geometric weighting emphasizing the latest data. In all the models we use a geometrically decreasing weighting with a common ratio 0.98 favoring the latest terms in the data sequences. For the case where we fit both the initial and the reopen phases, each phase is applied with this weighting independently, making sure that the end of the initial phase is fitted relatively close to the reported data. The data was obtained from multiple sources (*24, 29*)

We first illustrate the advantage of using the SIR-SD model over the traditional SIR model.

##### 5.2.1 SIR-SD model comparison and advantages

###### Discussion of Program 1

We first compared the SIR-SD model against the traditional SIR model where the transmission parameter *β* is a constant. To eliminate the influence of the release in the reopen phase we limit the fitting within only the initial phase before the reopen phase. We set *θ*_1_ = 0.9, *θ*_2_ = 0.05 in the loss function *L*(), so as to focus on fitting *G* instead of *D* and *I*_*H*_, in order to compare to the fitting of the traditional SIR model that only fits *G*. The results for our curve fitting are illustrated in Figure 3. The range specified to the optimizer for the parameter *γ* for SIR model was [0.04, 0.08] as the mean infectious period is reported to be within this range (*30*). In the SIR-SD model we used

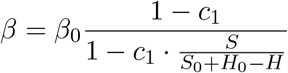

Note that before entering the reopen phase,

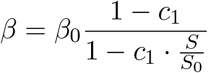

The SIR-SD model had a better fit than the SIR model, as illustrated in Figure 7 and as measured by RMSE values in Table 1.

**Figure 7:**
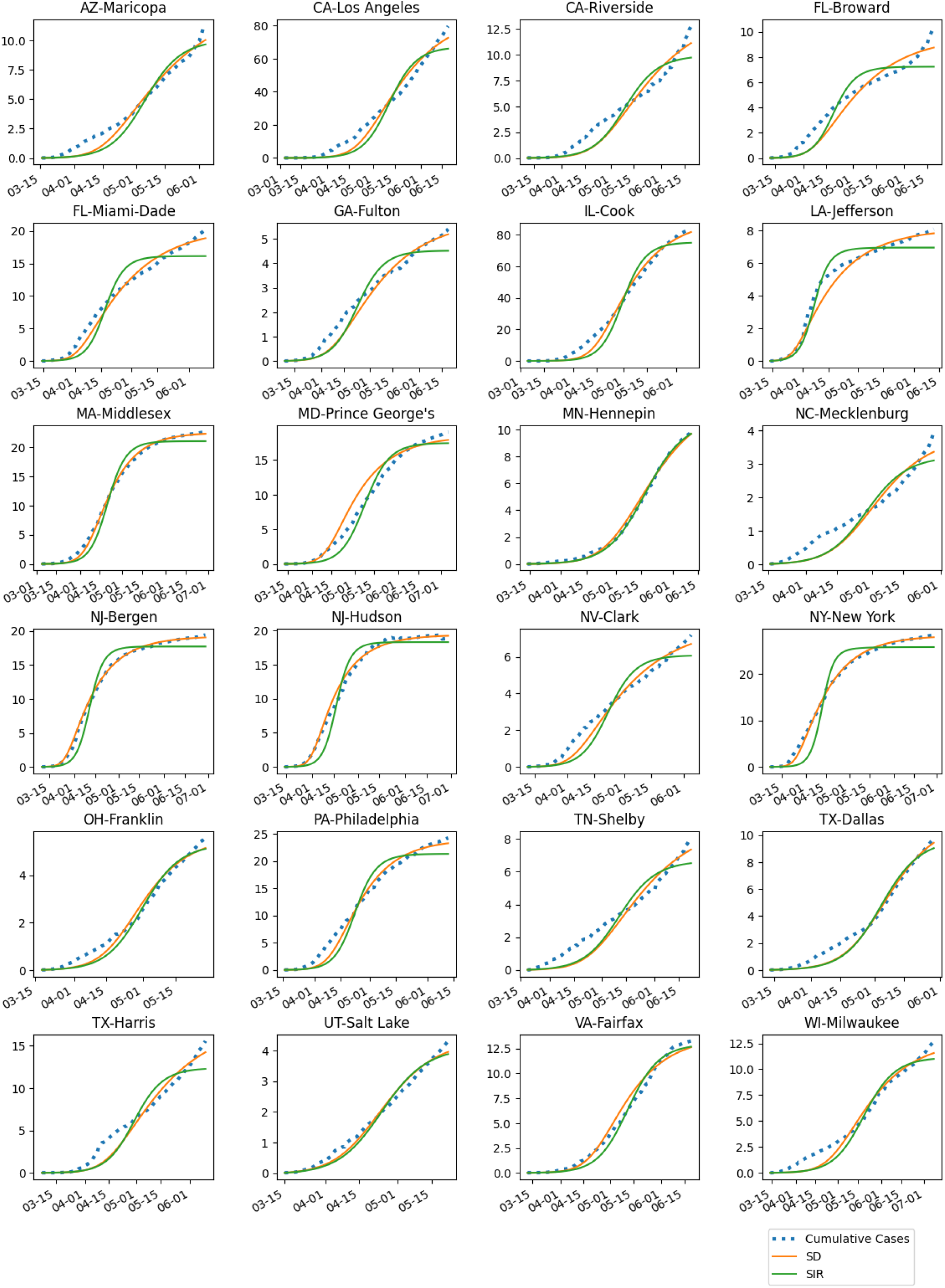
SIR-SD (SD in legend) provides a better fit for cumulative cases than the standard SIR model. Model fitting is for 24 US counties.

**Figure 8:**
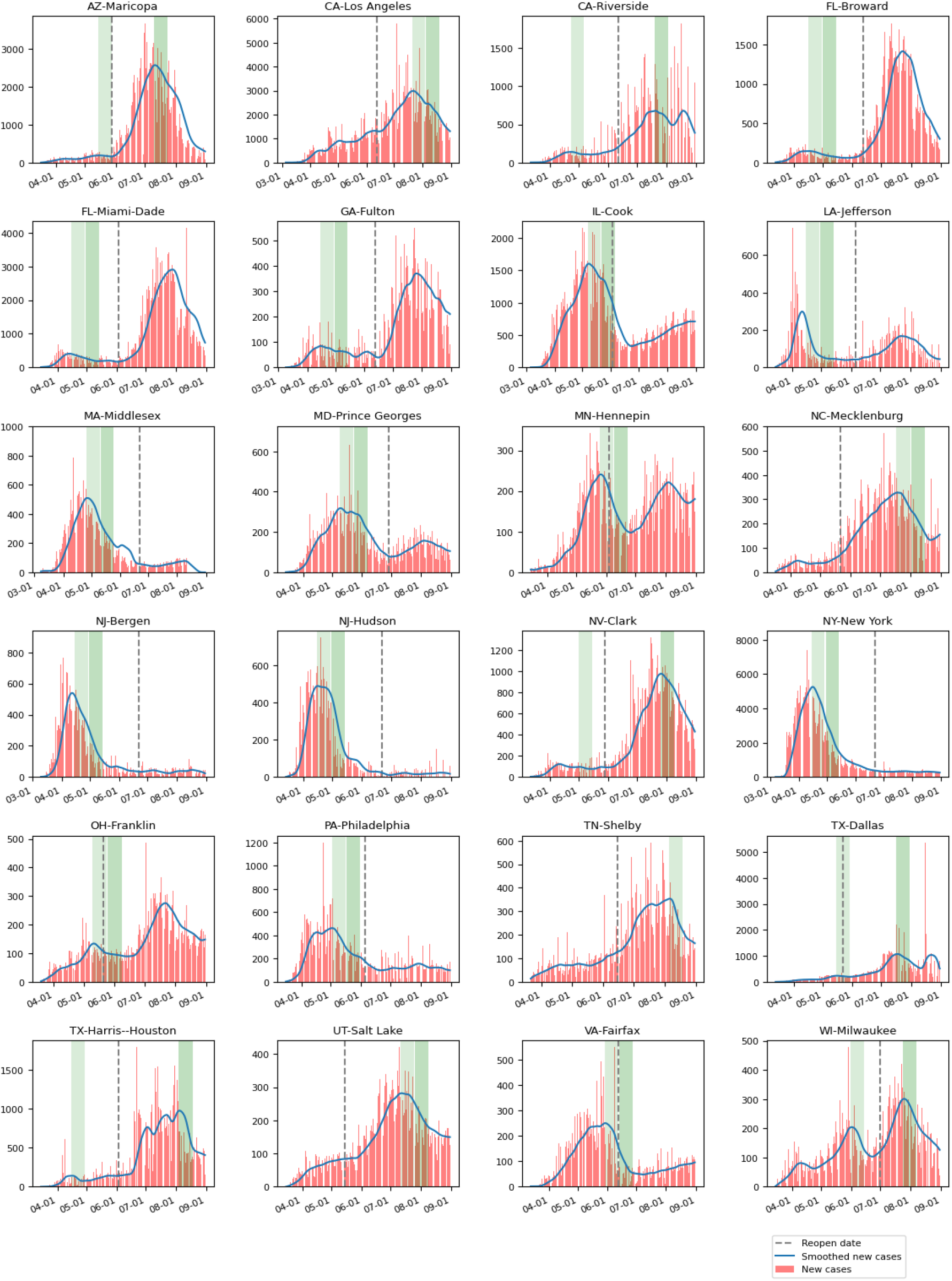
Policy Difference: Two-phase rule where reopen should be after second green band vs actual reopen date (dashed vertical line) for each county.

##### 5.2.2 SIR-SD with linear release Model fitting

###### Discussion of Program 2

To fit the model parameters during the lockdown phase and subsequently the re-opening phase we partition our fitting into two phases, the initial and the reopen phase. As mentioned before, each phase is weighted geometrically favoring the latest data. After some experimentation, we choose *θ*_1_ = 0.45, *θ*_2_ = 0.45 to weight different series, *G, D* and *I*_*H*_. In the reopen phase, in addition to the regular interactions between different compartments, we estimated a size of population, previously in hiding, that is to be released into the susceptible set, as well as the daily speed of that release, assuming a linear release function. It is worth mentioning that during our study we observed a drop in hospitalization and deaths in the reopening phase. In order to capture this we assigned multiplier parameters, *k* and *k*_2_ valued from 0 to 1 to the parameters *γ, α*_2_ in the reopening phase to modify the behavior of hospitalization and fatality. Table 4 shows the weighted RMSE and weighted R^2^ values of cumulative cases *G* and cumulative deaths *D* that measures our fitting method. After obtaining the best-fit parameters, we determined the relative standard deviation of the reported cases and deaths *G, D*, against our simulated data *G, D*. Monte Carlo simulations (1000 runs) were used applying random scatter via a Gaussian distribution using the standard deviation computed to obtain 95% confidence intervals of each day of the fitting. Using Monte Carlo simulations we also obtained the distributions of all the parameters. The key parameters and their 95% confidence intervals are provided in Table 5. The range of fitting produced and the model parameters were used to create the interval bands for the total number of cases, *G*, and deaths, *D*, illustrated by the shaded intervals in Figure 3.

**Table 4:**
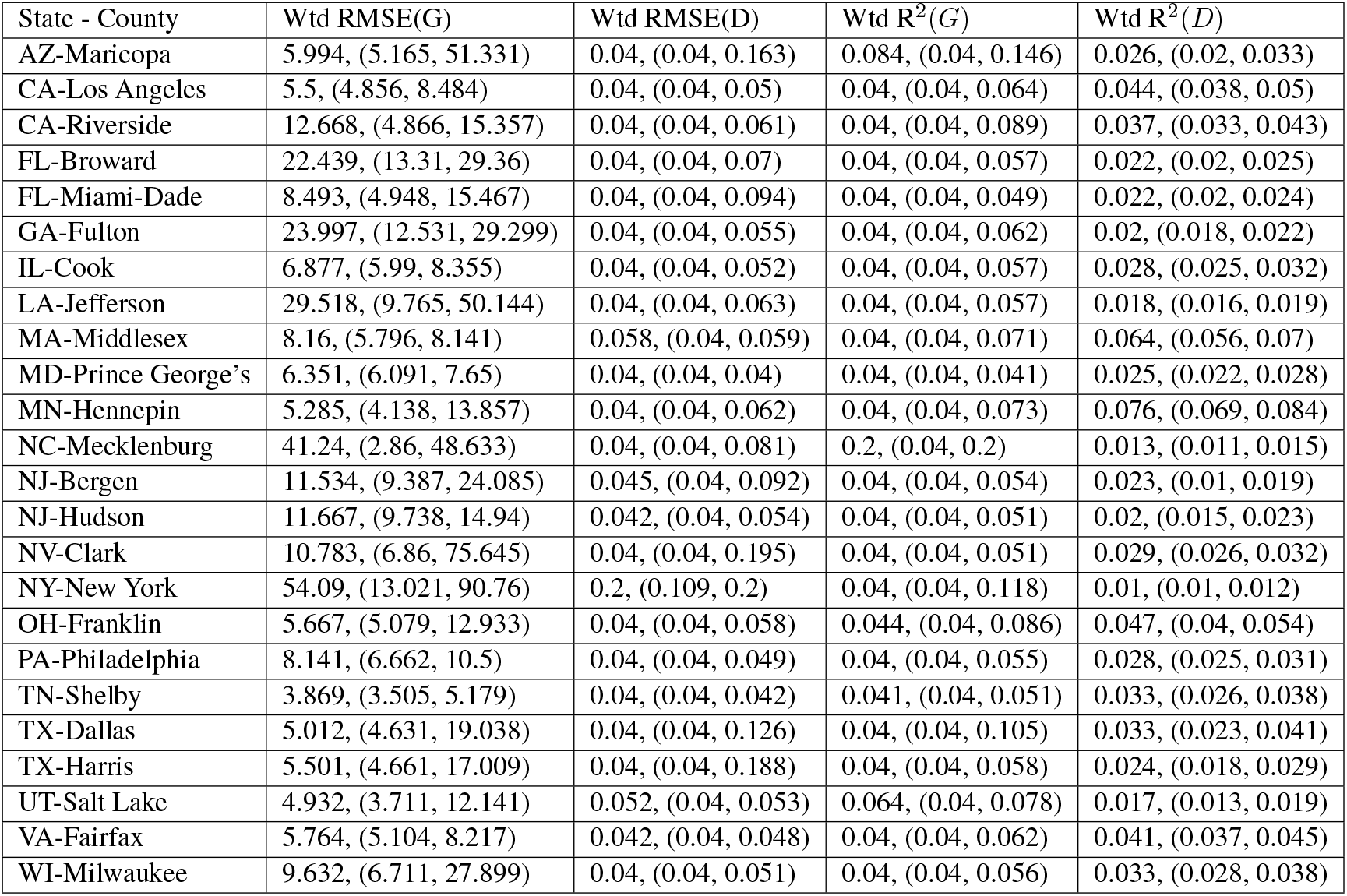
Weighted RMSE and R^2^ for combined fitting of cumulative infections (G) and deaths (D) over entire time range using SIR-SD-L model

| State - County | Wtd RMSE($G$) | Wtd RMSE($D$) | Wtd $R^2$($G$) | Wtd $R^2$($D$) |
| --- | --- | --- | --- | --- |
| AZ-Maricopa | 5.994, (5.165, 51.331) | 0.04, (0.04, 0.163) | 0.084, (0.04, 0.146) | 0.026, (0.02, 0.033) |
| CA-Los Angeles | 5.5, (4.856, 8.484) | 0.04, (0.04, 0.05) | 0.04, (0.04, 0.064) | 0.044, (0.038, 0.05) |
| CA-Riverside | 12.668, (4.866, 15.357) | 0.04, (0.04, 0.061) | 0.04, (0.04, 0.089) | 0.037, (0.033, 0.043) |
| FL-Broward | 22.439, (13.31, 29.36) | 0.04, (0.04, 0.07) | 0.04, (0.04, 0.057) | 0.022, (0.02, 0.025) |
| FL-Miami-Dade | 8.493, (4.948, 15.467) | 0.04, (0.04, 0.094) | 0.04, (0.04, 0.049) | 0.022, (0.02, 0.024) |
| GA-Fulton | 23.997, (12.531, 29.299) | 0.04, (0.04, 0.055) | 0.04, (0.04, 0.062) | 0.02, (0.018, 0.022) |
| IL-Cook | 6.877, (5.99, 8.355) | 0.04, (0.04, 0.052) | 0.04, (0.04, 0.057) | 0.028, (0.025, 0.032) |
| LA-Jefferson | 29.518, (9.765, 50.144) | 0.04, (0.04, 0.063) | 0.04, (0.04, 0.057) | 0.018, (0.016, 0.019) |
| MA-Middlesex | 8.16, (5.796, 8.141) | 0.058, (0.04, 0.059) | 0.04, (0.04, 0.071) | 0.064, (0.056, 0.07) |
| MD-Prince George's | 6.351, (6.091, 7.65) | 0.04, (0.04, 0.04) | 0.04, (0.04, 0.041) | 0.025, (0.022, 0.028) |
| MN-Hennepin | 5.285, (4.138, 13.857) | 0.04, (0.04, 0.062) | 0.04, (0.04, 0.073) | 0.076, (0.069, 0.084) |
| NC-Mecklenburg | 41.24, (2.86, 48.633) | 0.04, (0.04, 0.081) | 0.2, (0.04, 0.2) | 0.013, (0.011, 0.015) |
| NJ-Bergen | 11.534, (9.387, 24.085) | 0.045, (0.04, 0.092) | 0.04, (0.04, 0.054) | 0.023, (0.01, 0.019) |
| NJ-Hudson | 11.667, (9.738, 14.94) | 0.042, (0.04, 0.054) | 0.04, (0.04, 0.051) | 0.02, (0.015, 0.023) |
| NV-Clark | 10.783, (6.86, 75.645) | 0.04, (0.04, 0.195) | 0.04, (0.04, 0.051) | 0.029, (0.026, 0.032) |
| NY-New York | 54.09, (13.021, 90.76) | 0.2, (0.109, 0.2) | 0.04, (0.04, 0.118) | 0.01, (0.01, 0.012) |
| OH-Franklin | 5.667, (5.079, 12.933) | 0.04, (0.04, 0.058) | 0.044, (0.04, 0.086) | 0.047, (0.04, 0.054) |
| PA-Philadelphia | 8.141, (6.662, 10.5) | 0.04, (0.04, 0.049) | 0.04, (0.04, 0.055) | 0.028, (0.025, 0.031) |
| TN-Shelby | 3.869, (3.505, 5.179) | 0.04, (0.04, 0.042) | 0.041, (0.04, 0.051) | 0.033, (0.026, 0.038) |
| TX-Dallas | 5.012, (4.631, 19.038) | 0.04, (0.04, 0.126) | 0.04, (0.04, 0.105) | 0.033, (0.023, 0.041) |
| TX-Harris | 5.501, (4.661, 17.009) | 0.04, (0.04, 0.188) | 0.04, (0.04, 0.058) | 0.024, (0.018, 0.029) |
| UT-Salt Lake | 4.932, (3.711, 12.141) | 0.052, (0.04, 0.053) | 0.064, (0.04, 0.078) | 0.017, (0.013, 0.019) |
| VA-Fairfax | 5.764, (5.104, 8.217) | 0.042, (0.04, 0.048) | 0.04, (0.04, 0.062) | 0.041, (0.037, 0.045) |
| WI-Milwaukee | 9.632, (6.711, 27.899) | 0.04, (0.04, 0.051) | 0.04, (0.04, 0.056) | 0.033, (0.028, 0.038) |

**Table 5:** SIR-SD-L model parameters and confidence intervals for 24 counties

| State - County | $\beta$ (Best-Fit, (95%CI) | $\gamma$ (Best-Fit, (95%CI) | $\gamma_2$ (Best-Fit, (95%CI) | $\alpha_2$ (Best-Fit, (95%CI) |
| --- | --- | --- | --- | --- |
| AZ-Maricopa | 6.018, (5.443, 52.531) | 0.04, (0.04, 0.191) | 0.083, (0.04, 0.161) | 0.026, (0.019, 0.035) |
| CA-Los Angeles | 5.498, (4.812, 8.674) | 0.04, (0.04, 0.05) | 0.04, (0.04, 0.063) | 0.044, (0.038, 0.05) |
| CA-Riverside | 13.503, (5.124, 16.961) | 0.04, (0.04, 0.057) | 0.04, (0.04, 0.088) | 0.038, (0.033, 0.044) |
| FL-Broward | 20.17, (11.244, 27.032) | 0.04, (0.04, 0.064) | 0.04, (0.04, 0.063) | 0.022, (0.02, 0.025) |
| FL-Miami-Dade | 8.053, (4.522, 10.805) | 0.04, (0.04, 0.089) | 0.04, (0.04, 0.049) | 0.021, (0.019, 0.023) |
| GA-Fulton | 23.355, (12.379, 28.223) | 0.04, (0.04, 0.052) | 0.04, (0.04, 0.057) | 0.02, (0.018, 0.022) |
| IL-Cook | 6.877, (5.994, 8.365) | 0.04, (0.04, 0.052) | 0.04, (0.04, 0.057) | 0.028, (0.025, 0.033) |
| LA-Jefferson | 29.518, (10.369, 47.966) | 0.04, (0.04, 0.062) | 0.04, (0.04, 0.055) | 0.018, (0.016, 0.019) |
| MA-Middlesex | 7.229, (6.131, 8.598) | 0.057, (0.04, 0.065) | 0.04, (0.04, 0.069) | 0.064, (0.057, 0.07) |
| MD-Prince Georges | 6.29, (5.975, 7.624) | 0.04, (0.04, 0.04) | 0.04, (0.04, 0.042) | 0.025, (0.022, 0.027) |
| MN-Hennepin | 5.285, (4.246, 12.545) | 0.04, (0.04, 0.062) | 0.04, (0.04, 0.07) | 0.076, (0.069, 0.084) |
| NC-Mecklenburg | 43.564, (2.774, 46.959) | 0.04, (0.04, 0.075) | 0.2, (0.04, 0.2) | 0.013, (0.011, 0.015) |
| NJ-Bergen | 10.758, (9.12, 17.603) | 0.054, (0.04, 0.078) | 0.04, (0.04, 0.064) | 0.022, (0.016, 0.025) |
| NJ-Hudson | 10.855, (9.927, 16.921) | 0.045, (0.04, 0.058) | 0.04, (0.04, 0.047) | 0.019, (0.012, 0.021) |
| NV-Clark | 10.811, (6.776, 75.831) | 0.04, (0.04, 0.196) | 0.04, (0.04, 0.052) | 0.029, (0.026, 0.033) |
| NY-New York | 13.849, (11.537, 26.656) | 0.063, (0.04, 0.082) | 0.04, (0.04, 0.075) | 0.037, (0.026, 0.043) |
| OH-Franklin | 5.666, (5.433, 13.538) | 0.04, (0.04, 0.054) | 0.044, (0.04, 0.076) | 0.046, (0.041, 0.053) |
| PA-Philadelphia | 8.118, (6.728, 10.405) | 0.04, (0.04, 0.05) | 0.04, (0.04, 0.057) | 0.028, (0.025, 0.031) |
| TN-Shelby | 3.566, (3.328, 4.735) | 0.04, (0.04, 0.04) | 0.04, (0.04, 0.044) | 0.031, (0.025, 0.034) |
| TX-Dallas | 5.039, (4.514, 20.719) | 0.04, (0.04, 0.124) | 0.04, (0.04, 0.102) | 0.026, (0.019, 0.033) |
| TX-Harris-Houston | 6.85, (4.488, 24.164) | 0.04, (0.04, 0.14) | 0.04, (0.04, 0.05) | 0.01, (0.01, 0.011) |
| UT-Salt Lake | 4.953, (3.689, 12.711) | 0.056, (0.04, 0.055) | 0.069, (0.04, 0.081) | 0.017, (0.013, 0.019) |
| VA-Fairfax | 5.762, (5.1, 8.241) | 0.042, (0.04, 0.047) | 0.04, (0.04, 0.061) | 0.041, (0.038, 0.044) |
| WI-Milwaukee | 9.602, (6.576, 29.419) | 0.04, (0.04, 0.049) | 0.04, (0.04, 0.059) | 0.031, (0.027, 0.037) |

##### 5.2.3 Extended Simulation

We finally ran experiments to delay the release by 1 month to determine the decrease in cumulative cases by the end of September. We used the parameters determined by the model-fitting process. As shown in Figure 6, a one month delay contributes to a significant decrease in cumulative cases. The shaded area represents the cumulative cases for the actual release and the blue curve represents the cumulative cases if the release is delay by 1 month. In Table 3 we list out the ratio of the net increase in cumulative cases if we delay the reopen by 1 month, relative to net increase of the actually reported new cases, over the same time period. This period is over the range of dates starting from each county’s reopen date up to the end of August 2020. We see an average drop of 42% across the 24 counties.

###### Data Availability and Computer Code

All data is available at public sources (*24, 29*). All computer code is available from the authors and will be made publicly available

## Notes

### Competing Interest Statement

The authors have declared no competing interest.

### Funding Statement

Funded by NSF grant No. 2028274.

### Author Declarations

Approval provided by IRB Illinois Institute of Technology: Exempt Research. Exemption Category: Category 4. Secondary research for which consent is not required. (ii) Information, which may include information about biospecimens, is recorded by the investigator in such a manner that the identity of the human subjects cannot readily be ascertained directly or through identifiers linked to the subjects, the investigator does not contact the subjects, and the investigator will not re-identify subjects;

